# Histopathological assessments reveal retinal vascular changes, inflammation and gliosis in patients with lethal COVID-19

**DOI:** 10.1101/2021.02.25.21251531

**Authors:** Vijay K. Jidigam, Rupesh Singh, Julia C. Batoki, Caroline Milliner, Onkar B. Sawant, Vera L. Bonilha, Sujata Rao

## Abstract

**Purpose:** To assess for histopathological changes within the retina and the choroid and determine the long-term sequelae of the SARS-CoV-2 infection.

**Design:** Comparative analysis of human eyes.

**Subjects:** Eleven donor eyes from COVID-19 positive donors and similar age-matched donor eyes from patients with a negative test for SARS-CoV-2 were assessed.

**Methods:** Globes were evaluated ex-vivo with macroscopic, SLO and OCT imaging. Macula and peripheral regions were processed for epon-embedding and immunocytochemistry

**Main Outcome Measures:** Retinal thickness and histopathology, detection of SARS-CoV-2 Spike protein, changes in vascular density, gliosis, and degree of inflammation.

**Results:** Fundus analysis shows hemorrhagic spots and increased vitreous debris in several of the COVID-19 eyes compared to the control. OCT based measurements indicated an increased trend in retinal thickness in the COVID-19 eyes, however the difference was not statistically significant. Histology of the retina showed presence of hemorrhages and central cystoid degeneration in several of the donors. Whole mount analysis of the retina labeled with markers showed changes in retinal microvasculature, increased inflammation, and gliosis in the COVID-19 eyes compared to the controls. The choroidal vasculature displayed localized changes in density and signs of increased inflammation in the COVID-19 samples.

**Conclusions:** *In situ* analysis of the retinal tissue suggested that there are severe subclinical abnormalities that could be detected in the COVID-19 eyes. This study provides a rationale for evaluating the ocular physiology of patients that have recovered from COVID-19 infections to further understand the long-term effects caused by this virus.

## INTRODUCTION

We are amid the human coronavirus disease 2019 (COVID-19) pandemic, caused by severe acute respiratory syndrome coronavirus (SARS-CoV-2), which is of historic proportions, the likes of which we have not seen in 102 years. With >25 million cases confirmed, >490K deaths in the US (WHO COVID-19 Dashboard) it is one of the deadliest events in US history, and rates continuing to rise, the end is not in near sight. Despite being primarily a respiratory virus, COVID-19 can also present with non-respiratory signs, including ocular symptoms as conjunctival hyperemia, chemosis, epiphora, increased secretions, ocular pain, photophobia, and dry eye^1-6^. SARS-CoV-2 requires host cellular receptors (such as ACE2) for successful replication during infections. Immuno-histochemical studies and single-cell RNA-sequencing datasets have revealed both extra- and intra-ocular localization of SARS-CoV-2 receptors ACE2 receptor, and TMPRSS2 protease in human eyes^1,7-9^. The virus has also been detected within the anterior chamber and in the ocular fluids suggesting that ocular tissue may be directly affected due to Sars-CoV-2 infection^1,10,11^. Evidence of posterior eye involvement in SARS-COV-2 infection is still scarce, though some recent optical coherence tomography angiography (OCTA) based findings show that retinal microvasculature is affected in patients that recovered from COVID-19 infection^12,13^. However, a detailed histopathological analysis of the retinal tissue and the impact of the SARS-CoV-2 infection on ocular health and function has not been examined. Here we report a comprehensive analysis of eyes from post-mortem patients infected with the SARS-CoV-2 virus. Fundus imaging provides evidence of increased hemorrhage in the eyes of COVID-19 patients. In some SARS-CoV-2 positive patients, there was evidence of vascular anomalies consistent with ocular vein occlusion and reduced capillary density. Additionally, there is an overall increase in the number of microglial cells within the retina of COVID-19 patients. The microglial cells display abnormal morphological features indicative of microglial dystrophy. There is also increased gliosis in the COVID-19 eyes compared to the eyes from the age-matched control donors. To our knowledge, this is the first study that provides in vivo molecular information concerning the changes occurring within the retinal tissue of COVID-19 patients. We are still in the midst of the coronavirus outbreak and there is constantly emerging information regarding its long-term sequelae on various systems in the body. The data reported here provide a rationale for longitudinal ocular assessments in recovered patients to truly gain insights into understanding the long-term effects caused by this virus.

## METHODS

### Tissue acquisition and fixation

Donor eyes were obtained through Eversight (Cleveland, OH). Eye bank records accompanying the donor eyes indicated whether the donor had COVID-19. Pathology analysis was performed with the approval of the Cleveland Clinic Institutional Review Board (IRB #20-755) and Institutional Biosafety Committee (IBC# 2018). The research adhered to the tenets of the Declaration of Helsinki. Tissue from eleven COVID-19 donor eyes from seven donors was analyzed. Additional information about the donors is provided in Table 1 (Fig 1). Cornea and anterior segment analysis for the COVID-19 donors were recently reported by Sawant et al.^11^ For analysis, globes without the cornea were fixed and kept in 4% paraformaldehyde and 0.5% glutaraldehyde made in Dulbecco’s Phosphate Buffered Saline (D-PBS) buffer for at least a month.

**Table 1.** Human Donor Information.

| CASE | Age Range<sup>b</sup> | Gender<sup>c</sup> | Race<sup>d</sup> | PMI<sup>e</sup> | SARS-CoV-2 Testing |
| --- | --- | --- | --- | --- | --- |
| N<sup>a</sup> |  |  |  |  |  |
| 2 | 80-90 | F | C | 12 | Positive<sup>f</sup> |
| 4 | 60-65 | M | SA | 7 | Positive<sup>f</sup> |
| 5 | 70-75 | F | C | 27 | Positive<sup>f</sup> |
| 6 | 65-70 | M | H | 20 | Positive<sup>f</sup> |
| 7 | 55-60 | F | C | 11 | Positive<sup>f</sup> |
| 8 | 80-90 | F | H | 16 | Positive<sup>f</sup> |
| 10 | 45-50 | M | H | 13 | Positive<sup>f</sup> |
| 11 | 75-80 | M | C | 5 | Negative |
| 12 | 35-40 | F | C | 17 | Negative |
| 13 | 70-75 | M | C | 7 | Negative |
| 15 | 80-90 | M | C | 24 | Negative |
| 16 | 60-65 | F | C | 13 | Negative |
| 17 | 60-65 | M | C | 7 | Negative |

<sup>a</sup> Cornea and anterior segments analysis were reported in Sawant et al., *Ocul Surf.* 2020 Nov 8;S1542-0124(20)30168-3. doi: 10.1016/j.jtos.2020.11.002.
<sup>b</sup> Age: age at death (years);
<sup>c</sup> Gender: M = male, F= female;
<sup>d</sup> Race: AA = African American, C = caucasian, H = hispanic, SA = south asian
<sup>e</sup> Interval from death to preservation (hrs).
<sup>f</sup> Results reported in Sawant et al., *Ocul Surf.* 2020 Nov 8;S1542-0124(20)30168-3. doi: 10.1016/j.jtos.2020.11.002.

**Figure 1.**
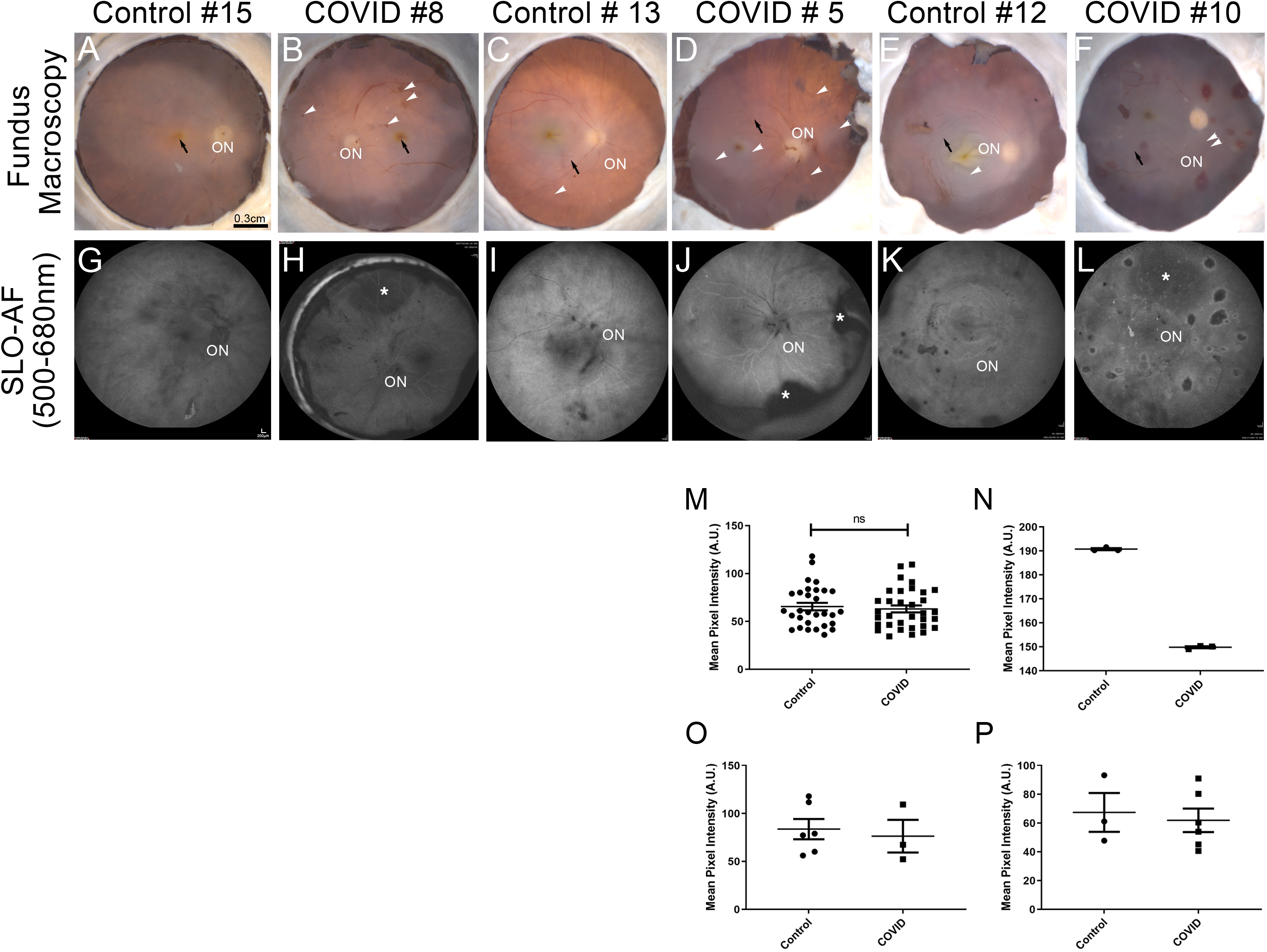
Ex-vivo imaging of COVID-19 donor eyes. Representative fundus (**A-F**) and SLO images (**G-L**) collected from COVID-19 and an age-similar controls. Fovea (black arrow) and optic nerve head (ON) were visible in all eyes. Hemorrhage spots (white arrowheads) were visible in most of the COVID-19 eyes. BAF images of COVID-19 (**H, J, L**) and control (**G, I, K**) eyes revealed a pattern that matched the fundus images. Also, in the COVID-19 eyes, the detached retinas were apparent with SLO (**H, J, L**, *). Mean intensity calculation using BAF SLO images with all age groups (**M**), age group 80-90 (**N**), age group 70-75 (**O**), and age group < 45 (**P**). Scale bar A-F= 0.3cm, G-L= 200μm.

### Ex-vivo imaging of globes

Globes were cut through the ora serrata, posterior poles were transferred to a chamber filled with D-PBS solution and imaged as previously described^14^. Briefly, fundus macrophotography (FM) images were collected using a Zeiss AxioCam MRC5 camera equipped with a macro zoom lens and AxioVision AC Software (Zeiss). Fundus autofluorescence was obtained using Blue Autofluorescence (BAF) mode of Heidelberg Spectralis confocal scanning laser ophthalmoscopy (SLO) (Heidelberg Engineering, Inc.). Optical Coherence Tomography images (OCT) were collected using spectral domain OCT system (Envisu R2210 UHR Leica Microsystems Inc.).

### Histopathology

Fragments of retina-RPE-choroid were cut from the periphery to the optic nerve head and placed in 2.5% glutaraldehyde in 0.1M cacodylate buffer, sequentially dehydrated in ethanol and embedded in Epon as previously reported ^15^. For light microscopy, semi-thin sections were cut with a diamond histotech knife, dried, and stained with toluidine blue. Slides were photographed with a Zeiss AxioImager.Z1 light microscope and the images were digitized using a Zeiss AxioCam MRc5 camera.

### Retinal flatmount and immunohistochemistry

A piece of retina and choroid were cut from the posterior pole of each eyecup. The retina was dissected and washed overnight in TBS. The RPE/choroid was incubated in the disodium salt of ethylenediaminetetraacetic acid for 1.5 hours and RPE was removed using a pipette before washing with TBS overnight and wholemount staining was performed as described with the exception that the tissues were incubated with the antibodies and the UEA lectin for 4 days^16^. Retinas were stained with chicken anti-glial fibrillary acidic protein (GFAP; 1:500; Millipore, Burlington, MA, USA), mouse anti-SARS-CoV S Protein (1:100; NR-614, BEI Resources, NIAID, Manassas, VA, USA), rabbit anti-Iba-1 (1:250; Wako Chemicals USA, Inc., Richmond, VA) and Ulex europaeus agglutinin-FITC (UEA lectin1:100; Genetex, Irvine, CA, USA). Choroids were stained with UEA Lectin and Iba-1 antibody. The submacular regions of the retina were imaged using a Leica confocal microscope. Retinal images were acquired from three different zones, one near the ONH and Vein, one near the middle and one towards the periphery^17,18^. All care was taken to ensure that similar regions were represented in the images. The choroids were imaged with Bruch’s membrane proximal to the objective.

## RESULTS

### Imaging of posterior globes

The combination of the fundus, confocal scanning laser ophthalmoscopy and optical coherence tomography imaging systems can provide a comprehensive characterization of retinal lesions before histopathology^19^. Therefore, these imaging techniques were performed on all COVID-19 and control eye donors; images obtained were qualitatively compared (Fig 1). Anatomical landmarks such as the optic nerve (ON) and fovea (Fig 1, black arrows) were identified in all donor’s eyes using all three imaging modalities. Fundus images displayed differences between the eyes from both groups, which included several hemorrhage spots in the COVID-19 eyes (Fig 1A to 1F, white arrowheads). SLO BAF imaging revealed detached retina areas in the COVID-19 eyes (Fig 1H, 1JI, and 1L, asterisks); further studies are needed to understand the cause of these retinal detachment areas. In addition, hemorrhage spots in the COVID-19 eyes were observed as hypofluorescent spots (Fig 1H, 1JI, and 1L, white arrowheads). Quantification of SLO images showed decreased BAF signal in the COVID-19 eyes compared to controls, but the increase was not significant (Fig 1M to 1P). Finally, OCT based measures for retinal thickness and optic nerve head depth showed slight increase in the COVID-19 eyes compared to controls, but the increase was not significant (Supplemental Fig S1). In general, COVID-19 posterior poles had more vitreous debris, most likely due to detached epiretinal membranes or cellular floaters.

### Histopathological and immunohistological findings in the retina of COVID-19 patients

Little is known about the ocular implications of COVID-19 disease. To gain insight into the disease’s retinal pathology, semi-thin sections of epon-embedded tissue were analyzed and compared to matched controls. The control donors’ retinas displayed each of the usual retinal lamina and the RPE (Fig 2A). The COVID-19 donors’ retina displayed retinal edema compared to the control retinas (Fig 2B). Interestingly, a few control retinas displayed variable areas of cystoid change and atrophy in the retina’s far periphery (Fig 2C). However, cystoid changes and atrophies were observed in the central area of the COVID-19 donors (Fig 2D). In our cohort of COVID-19 samples, we also observed hemorrhages of various sizes in the outer plexiform layer around the optic nerve head in a few COVID-19 donors (Fig 2E and F, asterisks); these were also autofluorescent in cryosections. The frequency of observed morphological findings detected in our cohort is provided in Table 2. Immunostaining of retinas with SARS-CoV-2 S protein antibody reacted with round cells within the retina of all the COVID-19 eyes close to the optic nerve head but not in the control retinas. However, in the retinas of two of the COVID-19 positive cases several cells were identified in the retina, choroid and optic nerve head (Fig 2H, and 2I, arrows) but not in the controls.

**Table 2.**
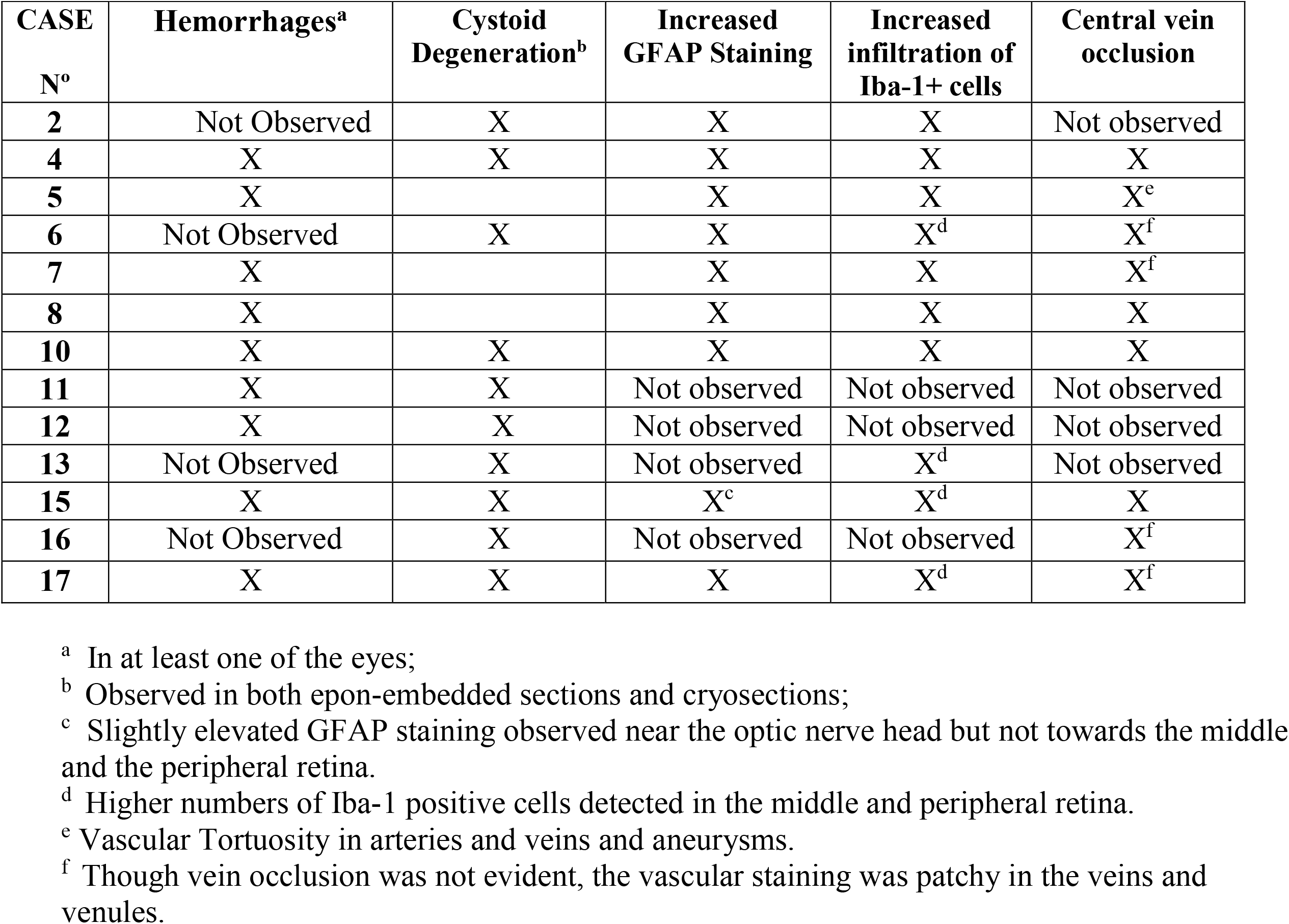
Assessment of retinal fundus and histological findings in donor eyes.

**Figure 2.**
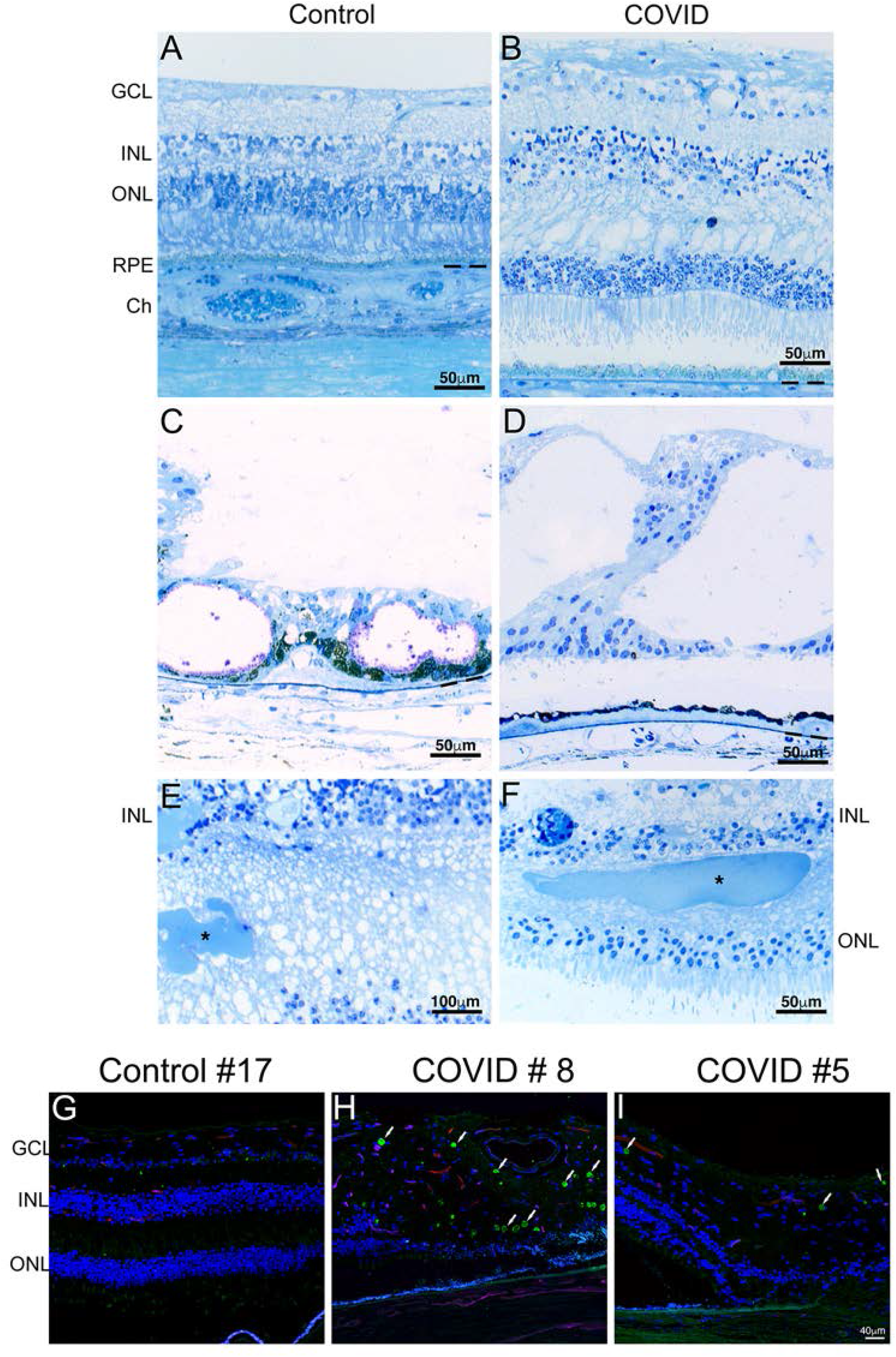
Histology and immunohistology of COVID-19 donor eyes. Representative toluidine blue-stained plastic 1 μm sections of retinas from COVID-19 donors (**B, D, E, F**) and an age-similar controls (**A, C**). Morphology of the control retina displayed typical retinal lamina. A few control retinas displayed cystoid degeneration (**C**) in the far periphery, while these were observed in the central retina of several COVID-19 eyes (**D**). Two of the COVID-19 retinas showed a cotton-wool exudate (**E, F**, asterisk) in the outer plexiform layer close to the optic nerve head. Immunofluorescence of two COVID-19 retinas labeled with antibodies to SARS-CoV S protein (**H-I**, Alexa488, green) showed the presence of several positive cells (arrows) when compared to control (**G**). GCL ganglion cell layer, INL inner nuclear layer, ONL outer nuclear layer, POS photoreceptor outer segments, RPE retinal pigment epithelium, Ch Choroid. Scale bar A-D, F= 50μm, E= 100μm, G-I= 40μm.

### Retinal microvasculature anomalies in COVID-19 patients

To gain insight into the disease’s retinal pathology, flatmounts of both retina and choroid were labeled with a lectin that labels the endothelium of vessels. In 4 out of 7 COVID-19 positive donor eyes, there were signs of major vein occlusion, indicated by constriction of the vein and increased signs of hemorrhages upstream of the constrictions (Supplemental Fig S2). Additionally, in the COVID-19 positive eyes, microvasculature density was severely reduced closer to the optic disc and around the veins (Fig 3A to 3F) compared to the age-matched control donor eyes. In several of the COVID-19 eyes, there was an observable loss of microvasculature and distinct thinning of the microcapillaries compared to the control eyes. In the choroidal plexus, vasculature did not appear to be different between the COVID-19 eyes and the age matched control cohorts. Though in some areas there was reduced focal lectin staining indicative of capillary dropouts unlike in the retinal vasculature, it was difficult to determine whether there is an overall reduction in the choroidal vasculature density in the COVID-19 patients (Supplemental Fig S3). Nevertheless, our analysis suggests that there are distinct histological changes in the retinal microvasculature. Though it is not possible to conclusively determine whether all reported vascular features can be completely attributed to the virus, based on the comparative analysis of the age-matched control donor eyes, it is important to note that other comorbidities cannot solely explain the observed differences.

**Figure 3.**
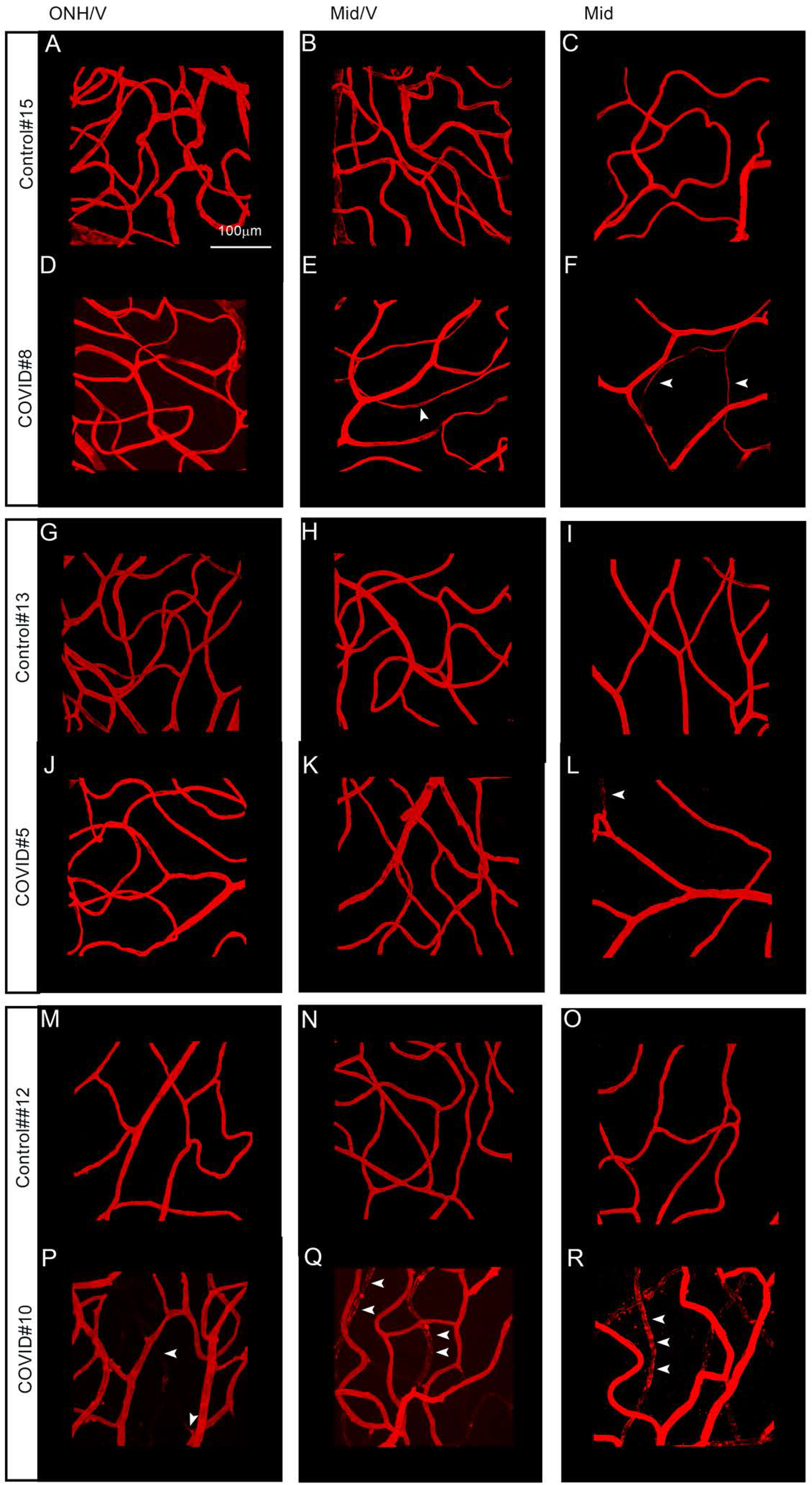
Retinal vascular abnormalities in COVID-19 patients. Representative images of retinal vasculature visualized using rhodamine-conjugated UEA lectin to label the blood vessels. (**A-R**). Representative images are from three different age groups. Age group 80-90 (**A-F**), Age group 70-75 (**G-L**) and age group <45 (M-R). To illustrate the spatial differences in vascular density of the retinal microvasculature, the retinal preparations were subdivided into three different zones, one near the optic nerve head closer to the vein (**A**,**D**,**G**,**J**,**M**,**P**) one near the middle closer to the vein (**B**,**E**,**H**,**K**,**N**,**Q**) and one between the middle and the periphery (**C**,**F**,**I**,**L**,**O**,**R**). Vessel density was severely reduced together with several capillaries showing signs of atrophy(white arrowheads) in the eyes from the COVID 19 patients (**E, F, L, R, Q**) compared to age matched controls (**B, C, I, O, N**). Most noticeable difference were observed in regions distal to the optic nerve head however in one of the COVID positive sample (**P-R**), regressing vessels could be detected in the entire retina. White arrowheads indicate the severe capillary dropout. ONH, Optic nerve head, V, Vein, Mid, Middle region, Scale bar: 100um.

### Gliosis and increased infiltration of microglial cells in the retina of COVID-19 patients

SARS-CoV-2 infection can lead to multisystem inflammatory syndrome. Ocular tissue, like many other tissues can be affected by this inflammatory process. To determine whether signs of increased inflammation can be detected, we assessed for gliosis and inflammation in the retina of COVID-19 patients. Retinal and choroidal preparations were examined using GFAP, a marker for astrocytes (Fig 4A to 4L) and Iba-1, to label the microglial cells (Fig 4A’ to 4L’). Irrespective of whether the eyes were from COVID-19 positive or negative individuals, Iba-1 positive microglial cells could be detected in all the eyes. As a result of aging, an increasing proportion of microglial cells display abnormal morphological features such as shortened, gnarled, beaded, or fragmented cytoplasmic processes, and loss of fine ramifications and formation of spheroidal swellings; these changes are designated collectively as microglial dystrophy^20^. These changes are determined to be different than what occurs during microglial activation which is defined as hypertrophic microglia^20-23^. In all the eyes examined, there was evidence for both hypertrophic and dystrophic microglia, though there was evidence of microglial dystrophy in more of the COVID-19 eyes compared to the controls. (Fig 4E’-4L’, hypertrophic are indicated with red arrow heads, while dystrophic features are indicated with blue and yellow arrowheads; magenta is normal morphology) Additionally, in all the COVID-19 eyes examined, there was also an overall increase in the number of microglial cells (Fig 4C’, 4D’, 4G’, 4H’, 4K’, 4L’). This increase was observed closer to the optic nerve head and towards the middle and the peripheral retina. An overall assessment of the increased microglial cells is also indicated in Table 2. Besides the differences in the numbers, microglial cells also displayed activated microglia characteristics with enlarged spheroidal morphology and retracted processes. Similar to our retinal observations, there was evidence of increased microglial cells in the choroid (Supplemental Fig S2). A direct or systemic inflammation can result in the activation of the astrocytes along with the microglial cells. Astrocyte network were visualized with GFAP. GFAP immunoreactivity was highly variable with temporal differences within macular and perimacular regions that were examined. In general, closer to the optic nerve, the GFAP immunoreactivity was increased in the COVID-19 patients compared to their age matched controls. The astrocyte networks also appear to be different, with more elongated cell processes closer to the ONH (Fig 4C, 4G,4K) but towards the mid and peripheral regions, these elongated bundles appear to overlap and form dense mesh like structures (Fig 4D,4H, 4L). The increased GFAP immunoreactivity suggests signs of increased gliosis within the retina of patients infected with SARS-CoV-2 virus. However, due to the limited number of eyes examined, it is difficult to conclusively assess the influences of other comorbidities such as age and sex for the analysis.

**Figure 4.**
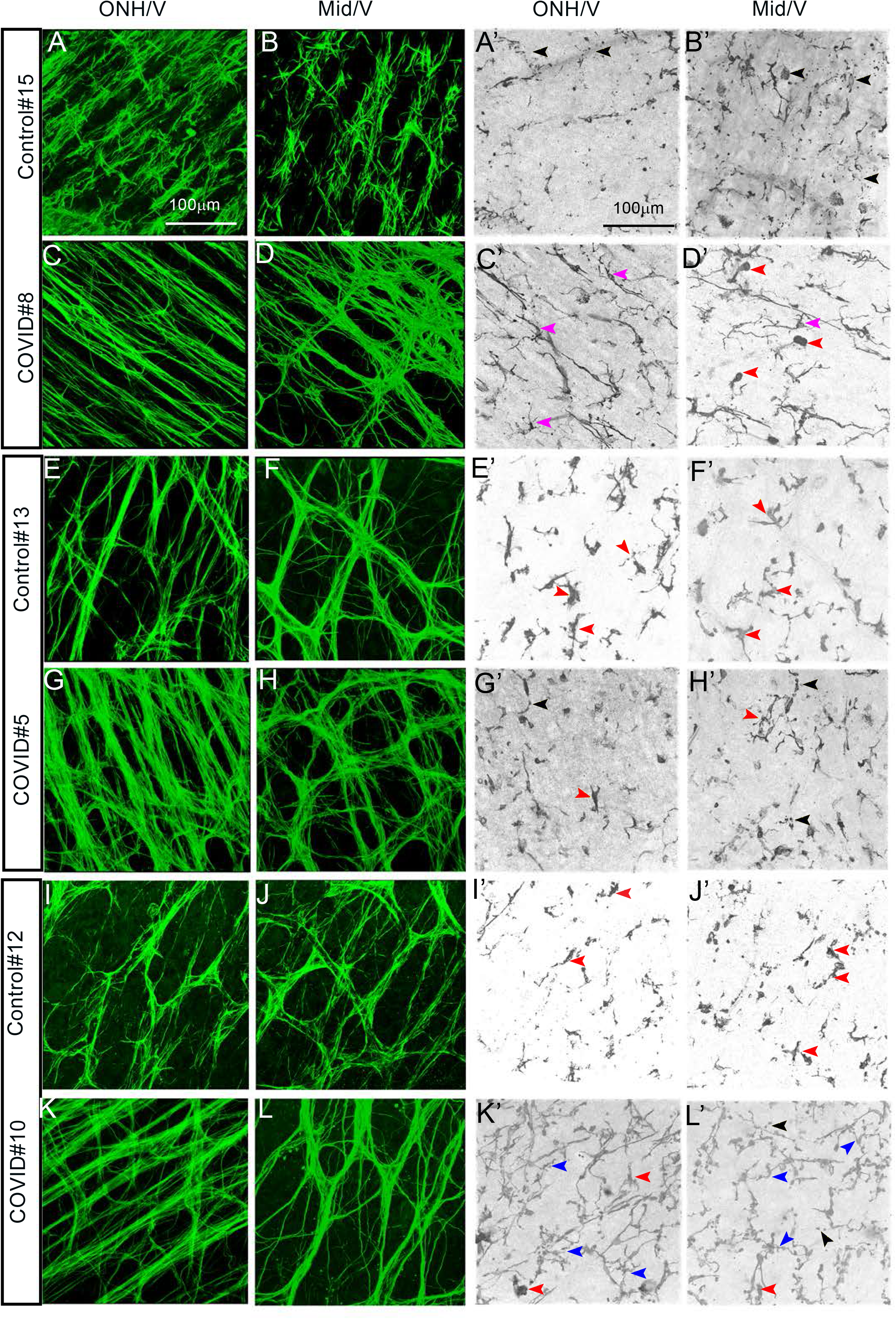
Retinal vascular anomalies are accompanied by gliosis and inflammation. Retinal whole mounts stained with an antibody for GFAP for glial cells (**A-L**; green) and Iba1 for microglia (**A’-L’**, grey). There is an overall increase in GFAP immunoreactivity near the ONH regions (**G**,**K**) in the COVID-19 patients when compared to age matched controls (**E, I**). However, in one COVID negative case (**A, B**) there is a similar increase in GFAP immunoreactivity near the ONH area (**A**), but not near the middle regions (**B**). In general the GFAP positive cells appear to form dense networks and overlapping cable like structures indicative of increased gliosis (**A’-L’**) Representative images of retinal tissues stained with Iba1to visualize the microglial cells. In all the COVID positive eyes, in the regions distal to the optic nerve and right next to the vein. (**C’, D’ G’ H’K’ L’**) there is an increase in the total number of microglial cells irrespective of the age of the deceased patient compared to the control eyes (**B’**,**F’**,**J’**). In general, in all of the eyes examined, several of the microglial cells displayed hypertrophic morphology (red arrowhead, **D’-L’**) with some showing fragmented processes (black arrowhead **A’, B’, G’, H’, L’**),beady, processes (blue arrowheads, **K’, L’**) indicative of dystrophic microglia with very few normal looking microglia only seen in one eye examined (Magenta arrowheads, **C’**,**D’**). ONH, Optic nerve head, V, Vein, Mid, Middle region, Scale bar: 100um.

## DISCUSSION

Though the SARS-CoV-2 is primarily a respiratory tract virus, there is sufficient evidence that the virus can be detected in several other tissues like the peripheral and central nervous systems. The presence of the ACE2 receptor and the cofactor TMPRSS2 in several anatomical parts of the anterior surface suggest that the SAR-CoV-2 infection of the eye tissues, especially at the limbus and the cornea is possible. In agreement with this observation, a recent report estimated the prevalence of ocular manifestations in COVID-19 patients to range between 2 to 32% ^24^. The ophthalmic manifestations appear to be associated with the disease severity of COVID-19 ^1,25^. Several groups have reported varying viral RNA and protein levels within the tears, conjunctiva, cornea, and vitreous^11,26^. The presence of viral ribonucleic acid (RNA) of SARS-CoV-2 was reported in three of fourteen human retinas of deceased patients with confirmed COVID-19 by real-time reverse transcriptase-polymerase chain reaction (RT-PCR)^27^. In this study, SARS-CoV-2 S protein immunoreactivity showed different degrees of distinct and specific localization in round cells within the retina of all the COVID-19 eyes close to the optic nerve head. The anterior segments of our COVID-19 cohort’s eyes were previously analyzed^11^. Among eleven eyes recovered from seven COVID-19 donors: three conjunctival, one anterior corneal, five posterior corneal, and three vitreous swabs tested positive for SARS-CoV-2 RNA. Cases of SARS-CoV-2 have been known to induce anterior segment pathologies such as conjunctivitis and anterior uveitis and posterior pathologies, including retinitis, optic neuritis, choroiditis with retinal detachment and retinal vasculitis^25,28^. These studies further illustrate the importance of long-term assessments of ocular physiology of individuals that have recovered from COVID-19.

To date, there is no detailed cellular and molecular characterization of the retina in SARS-CoV-2 infected patients. Previous studies reported retinal lesions in outpatients after confirmed SARS-CoV-2 infection with mild to moderate symptoms. Findings included non-specific and controversial hyper-reflective OCT lesions in the ganglion cell and inner plexiform layers, microhemorrhages, and nerve fiber infarcts^29^. Other studies identified hemorrhages, cotton wool spots, dilated veins and tortuous vessels in fundus photographs^13,30^. These are commonly seen as manifestations of diabetes mellitus and systemic hypertension but are also associated with several other etiologies, including ischemic, embolic, connective tissue, neoplastic, and infectious ^31-33^. CWSs compromise localized accumulations of axoplasmic debris at the level of retinal ganglion cell axons resulting from axoplasmic flow interruption due to vascular or mechanical causes ^34^. In our histological study, we detected amorphous debris in the outer plexiform layer corresponding to hemorrhages. Another exciting finding observed in COVID-19 eyes was the presence of a large number of cystoid lesions spread across the entire retina that closely resemble the retinal alterations of patients suffering from cystoid macular edema (CME)^35,36^ is observed in human retinal diseases like AMD, diabetic retinopathy, retinal vein occlusion, retinitis pigmentosa^35,37^, optic atrophy^38^, among others^36,39^. The presence of cysts causes a thickening of the affected retina and decreases visual acuity. Moreover, the neuroretina’s compression, the nerve fibers and capillaries by the cystic alterations further contribute to retinal degeneration and aggravation of hypoxic conditions.

Though, data on histopathological analysis of retinal vasculature and choriocapillaris is scarce, there have been a few studies on OCTA based findings in patients with SARS-Cov-2. In a recent study by Abrishami et al,^12^ OCTA was performed 2 weeks after recovery from systemic COVID-19 and mean vessel density in the superficial and deep plexus were significantly reduced in the COVID-19 cohort versus the age matched controls. In another study by Turker et al^40^, a reduction in retinal vessel density of the superficial and deep capillary plexus was reported. In their study, the measurements were done within one week of discharge after complete recovery. Another OCTA based study reported several retinal findings including hemorrhages, cotton wool spots, dilated veins, tortuous vessel and changes in mean arterial and vein diameter in patients with COVID-19^13^. Although the authors stated that such findings indicating microangiopathy might be secondary to COVID-19 or incidental, they also speculated that the virus itself or the systemic treatments used might have triggered microangiopathy in patients with systemic vascular disease. In the present study, all retinal and choroicapillaris changes were examined in post-mortem tissue. We detect similar reduction in retinal vascular density in COVID-19 patients along with increased vessel tortuosity, vein occlusions and hemorrhages. However, unlike the retinal vasculature, the choroidal vasculature did not appear to be severely affected as a result of the COVID-19 infection. It is important to note that there are studies that have found little or no change in retinal vascular density^41^. These apparent differences in reported observations, could be due to the disease severity, study populations, diagnostic criteria and methodologies used in the different studies. It still remains to be determined whether the changes in the retinal vasculature is due to a direct infection of the retina or whether these are part of a common systemic vascular diseases such as diabetes mellitus, chronic kidney disease and hypertension.

We also find evidence of increased microglial cells in the retina of the COVID-19 eyes. There is no reported evidence yet for increased infiltration of inflammatory cells in the retina, though a recent case study reported a possible association between COVID-19 and Papillophlebitis, a rare condition that occurs due to a consequence of inflammation of the retinal vein^42^. Chronological aging is associated with a significant increase in the total numbers of both hypertrophic as well as dystrophic microglia. A recent study showed that dystrophy are the disease associated microglia morphology^21^. In the present study we see evidence for increased microglial cells in the COVID-19 eyes and several of these appear to show the characteristic of microglial dystrophy and hypertrophy. Hypertrophic microglia are associated with increased microglial activation. However, due to the very low numbers of eyes examined and the subjectivity associated with classifying a hypertrophic, dystrophic and normal microglia purely based on morphological attributes, it is difficult to infer whether these dystrophic/hypertrophic morphological features is associated with changes in the microglia. While there is likely more than one explanation for the differences detected in the microglial morphology, whether this is a direct consequence of the SARS-CoV-2 infection remains to be determined. However, these preliminary findings are interesting and suggest that there could be an increase in the secretion of the pro-inflammatory molecules as a result of the microglial dystrophy and warrant further investigation. Though, there is no report on activation of microglia in the retina, there is some evidence from neuropathological findings in patients who have died from COVID-19 that in a significant number of these patients, various gliosis stages with diffuse activation of microglia and astrocytes could be detected. In our study we also find evidence of increased astrocyte activation as indicated by the increase in GFAP immunoreactivity. The astrocyte morphology also appears to be different with more elongated morphology closer to the ONH and dense mesh like networks toward the middle and the periphery. Such phenotypic heterogeneity has been associated with different responses of the astrocytes to an injury and their adaptive functions. Importantly, the dense mesh like network is indicative of scar formation after an injury^43,44^. GFAP has been also detected in the plasma of patients with COVID-19 and the amount is directly correlated with the severity of the disease^45,46^. Whether the observed activation is temporary and is resolved after the infection is gone, remains to be seen. However, even short-term increase in GFAP can be detrimental to the underlying neuronal cells and can result in focal damage. Despite the small sample size, the work presented here raises the possibility of subclinical vascular deficits combined with increased inflammation in patients with severe disease who have recovered from COVID-19 infection.

This study has some limitations, including the small sample size and the broad inclusion criteria. The severity of the viral infection is unknown, and the duration of hospitalization could severely impact the histopathological findings, thus limiting the generalization of the findings. Owing to the small numbers of available control eyes available, the cause of death for these control cohorts will notably have a huge impact on all the assessments reported. All care was taken to ensure that the investigated cohorts were closely matched in terms of age as well as the duration that these patients were maintained on ventilators. These limitations do not appear to change the results as many of the reported findings could only be observed in the deceased patients with COVID-19. Further evaluation with a much larger sample size is needed to determine the effects of SAR-CoV-2 infection on retinal morphology, vasculature, inflammation and gliosis.

In conclusion, we observed several ocular anomalies the COVID-19 cohorts compared to the control cohorts. Surprisingly, despite the small sample size, there were some consistent differences detected between the patient cohorts and the COVID-19 negative patients. Of note are the subclinical microvasculature features that we observed. As some of these observations have not been noted previously these histopathological analyses of the post-mortem eyes from the COVID-19 patients suggest that as more individuals recover from the COVID-19 infections, depending on the severity of their illness these individuals may present with ocular clinical symptoms that have not been examined previously. Therefore, a heightened vigilance for the long -term disease sequelae in other tissues like the eye is warranted.

## Data Availability

data will be available

## ACKNOWLEDGMENTS

This research was funded by grants from the U.S. National Institutes of Health/National Eye Institute EY027077-01 (S.R), RPB1503 (S.R.), EY027750 (VLB), a National Eye Institute P30-EY025585 Core Grant, a Cleveland Eye Bank Foundation Grant awarded to the Cole Eye Institute, and Research to Prevent Blindness Challenge Grant. Research activities at Eversight are supported by funding from LC Industries (Durham, NC), Eye Bank Association of America and Connecticut Lions Eye Research Foundation. The authors declare no conflicts of interest.

## AUTHORS CONTRIBUTIONS

VLB, and SR contributed to study conception and design, and accessed and verified the data. VKJ drafted the manuscript. All authors contributed to data acquisition, interpreted the data, critically revised the manuscript, and provided approval for the final version of the manuscript to be published.

## REFERENCES

1. Chen L, Deng C, Chen X, et al. Ocular manifestations and clinical characteristics of 534 cases of COVID-19 in China: A cross-sectional study. medRxiv. 2020:2020.2003.2012.20034678.

2. Abrishami M, Tohidinezhad F, Daneshvar R, et al. Ocular Manifestations of Hospitalized Patients with COVID-19 in Northeast of Iran. Ocul Immunol Inflamm. 2020;28(5):739–744.

3. Hong N, Yu W, Xia J, Shen Y, Yap M, Han W. Evaluation of ocular symptoms and tropism of SARS-CoV-2 in patients confirmed with COVID-19. Acta Ophthalmol. 2020.

4. Cheema M, Aghazadeh H, Nazarali S, et al. Keratoconjunctivitis as the initial medical presentation of the novel coronavirus disease 2019 (COVID-19). Can J Ophthalmol. 2020;55(4):e125–e129.

5. Yordi S, Ehlers JP. COVID-19 and the eye. Cleve Clin J Med. 2020.

6. Jin YP, Trope GE, El-Defrawy S, Liu EY, Buys YM. Ophthalmology-focused publications and findings on COVID-19: A systematic review. Eur J Ophthalmol. 2021:1120672121992949.

7. Zhou L, Xu Z, Castiglione GM, Soiberman US, Eberhart CG, Duh EJ. ACE2 and TMPRSS2 are expressed on the human ocular surface, suggesting susceptibility to SARS-CoV-2 infection. Ocul Surf. 2020;18(4):537–544.

8. Leonardi A, Rosani U, Brun P. Ocular Surface Expression of SARS-CoV-2 Receptors. Ocul Immunol Inflamm. 2020;28(5):735–738.

9. Wagner J, Jan Danser AH, Derkx FH, et al. Demonstration of renin mRNA, angiotensinogen mRNA, and angiotensin converting enzyme mRNA expression in the human eye: evidence for an intraocular renin-angiotensin system. Br J Ophthalmol. 1996;80(2):159–163.

10. Casagrande M, Fitzek A, Spitzer MS, et al. Presence of SARS-CoV-2 RNA in the Cornea of Viremic Patients With COVID-19. JAMA Ophthalmol. 2021.

11. Sawant OB, Singh S, Wright RE, 3rd, et al. Prevalence of SARS-CoV-2 in human post-mortem ocular tissues. Ocul Surf. 2021;19:322–329.

12. Abrishami M, Emamverdian Z, Shoeibi N, et al. Optical coherence tomography angiography analysis of the retina in patients recovered from COVID-19: a case-control study. Can J Ophthalmol. 2021;56(1):24–30.

13. Invernizzi A, Torre A, Parrulli S, et al. Retinal findings in patients with COVID-19: Results from the SERPICO-19 study. EClinicalMedicine. 2020;27:100550.

14. Bonilha VL, Rayborn ME, Bell BA, Marino MJ, Fishman GA, Hollyfield JG. Retinal Histopathology in Eyes from a Patient with Stargardt disease caused by Compound Heterozygous ABCA4 Mutations. Ophthalmic Genet. 2015:1–11.

15. Bonilha VL, Shadrach KG, Rayborn ME, et al. Retinal deimination and PAD2 levels in retinas from donors with age-related macular degeneration (AMD). Exp Eye Res. 2013;111:71–78.

16. Edwards MM, Bonilha VL, Bhutto IA, et al. Retinal Glial and Choroidal Vascular Pathology in Donors Clinically Diagnosed With Stargardt Disease. Invest Ophthalmol Vis Sci. 2020;61(8):27.

17. Stokoe NL, Turner RW. Normal retinal vascular pattern. Arteriovenous ratio as a measure of arterial calibre. Br J Ophthalmol. 1966;50(1):21–40.

18. Alam M, Toslak D, Lim JI, Yao X. Color Fundus Image Guided Artery-Vein Differentiation in Optical Coherence Tomography Angiography. Invest Ophthalmol Vis Sci. 2018;59(12):4953–4962.

19. Bagheri N, Bell BA, Bonilha VL, Hollyfield JG. Imaging human postmortem eyes with SLO and OCT. Adv Exp Med Biol. 2012;723:479–488.

20. Streit WJ, Sammons NW, Kuhns AJ, Sparks DL. Dystrophic microglia in the aging human brain. Glia. 2004;45(2):208–212.

21. Shahidehpour RK, Higdon RE, Crawford NG, et al. Dystrophic microglia are a disease associated microglia morphology in the human brain. bioRxiv. 2020:2020.2007.2030.228999.

22. Conde JR, Streit WJ. Microglia in the aging brain. J Neuropathol Exp Neurol. 2006;65(3):199–203.

23. Miller KR, Streit WJ. The effects of aging, injury and disease on microglial function: a case for cellular senescence. Neuron Glia Biol. 2007;3(3):245–253.

24. Ulhaq ZS, Soraya GV. The prevalence of ophthalmic manifestations in COVID-19 and the diagnostic value of ocular tissue/fluid. Graefes Arch Clin Exp Ophthalmol. 2020;258(6):1351–1352.

25. Wu P, Duan F, Luo C, et al. Characteristics of Ocular Findings of Patients With Coronavirus Disease 2019 (COVID-19) in Hubei Province, China. JAMA Ophthalmol. 2020;138(5):575–578.

26. Ocansey S, Abu EK, Abraham CH, et al. Ocular Symptoms of SARS-CoV-2: Indication of Possible Ocular Transmission or Viral Shedding. Ocul Immunol Inflamm. 2020;28(8):1269–1279.

27. Casagrande M, Fitzek A, Puschel K, et al. Detection of SARS-CoV-2 in Human Retinal Biopsies of Deceased COVID-19 Patients. Ocul Immunol Inflamm. 2020;28(5):721–725.

28. Seah I, Agrawal R. Can the Coronavirus Disease 2019 (COVID-19) Affect the Eyes? A Review of Coronaviruses and Ocular Implications in Humans and Animals. Ocul Immunol Inflamm. 2020;28(3):391–395.

29. Marinho PM, Marcos AAA, Romano AC, Nascimento H, Belfort R Jr., Retinal findings in patients with COVID-19. Lancet. 2020;395(10237):1610.

30. Pereira LA, Soares LCM, Nascimento PA, et al. Retinal findings in hospitalised patients with severe COVID-19. Br J Ophthalmol. 2020.

31. Mansour AM, Jampol LM, Logani S, Read J, Henderly D. Cotton-wool spots in acquired immunodeficiency syndrome compared with diabetes mellitus, systemic hypertension, and central retinal vein occlusion. Arch Ophthalmol. 1988;106(8):1074–1077.

32. Mansour AM, Jampol LM, Hrisomalos NF, Greenwald M. Case report. Cavernous hemangioma of the optic disc. Arch Ophthalmol. 1988;106(1):22.

33. Brown GC, Brown MM, Hiller T, Fischer D, Benson WE, Magargal LE. Cotton-wool spots. Retina. 1985;5(4):206–214.

34. McLeod D, Restori M, Wright JE. Rapid B-scanning of the vitreous. Br J Ophthalmol. 1977;61(7):437–445.

35. Scholl S, Kirchhof J, Augustin AJ. Pathophysiology of macular edema. Ophthalmologica. 2010;224 Suppl 1:8–15.

36. Rotsos TG, Moschos MM. Cystoid macular edema. Clin Ophthalmol. 2008;2(4):919–930.

37. Strong S, Liew G, Michaelides M. Retinitis pigmentosa-associated cystoid macular oedema: pathogenesis and avenues of intervention. Br J Ophthalmol. 2017;101(1):31–37.

38. De Bats F, Wolff B, Vasseur V, et al. “En-face” spectral-domain optical coherence tomography findings in multiple evanescent white dot syndrome. J Ophthalmol. 2014;2014:928028.

39. Zhang X, Zeng H, Bao S, Wang N, Gillies MC. Diabetic macular edema: new concepts in pathophysiology and treatment. Cell Biosci. 2014;4:27.

40. Turker IC, Dogan CU, Guven D, Kutucu OK, Gul C. Optical coherence tomography angiography findings in patients with COVID-19. Can J Ophthalmol. 2021.

41. Savastano MC, Gambini G, Cozzupoli GM, et al. Retinal capillary involvement in early post-COVID-19 patients: a healthy controlled study. Graefes Arch Clin Exp Ophthalmol. 2021.

42. Insausti-Garcia A, Reche-Sainz JA, Ruiz-Arranz C, Lopez Vazquez A, Ferro-Osuna M. Papillophlebitis in a COVID-19 patient: Inflammation and hypercoagulable state. Eur J Ophthalmol. 2020:1120672120947591.

43. Wanner IB, Anderson MA, Song B, et al. Glial scar borders are formed by newly proliferated, elongated astrocytes that interact to corral inflammatory and fibrotic cells via STAT3-dependent mechanisms after spinal cord injury. J Neurosci. 2013;33(31):12870–12886.

44. Burda JE, Sofroniew MV. Reactive gliosis and the multicellular response to CNS damage and disease. Neuron. 2014;81(2):229–248.

45. Kanberg N, Ashton NJ, Andersson LM, et al. Neurochemical evidence of astrocytic and neuronal injury commonly found in COVID-19. Neurology. 2020;95(12):e1754–e1759.

46. Virhammar J, Naas A, Fallmar D, et al. Biomarkers for central nervous system injury in cerebrospinal fluid are elevated in COVID-19 and associated with neurological symptoms and disease severity. Eur J Neurol. 2020.

