## SUPPLEMENTARY FIGURES for "Histopathological assessments reveal retinal vascular changes, inflammation and gliosis in patients with lethal COVID-19"

**Figure S1**

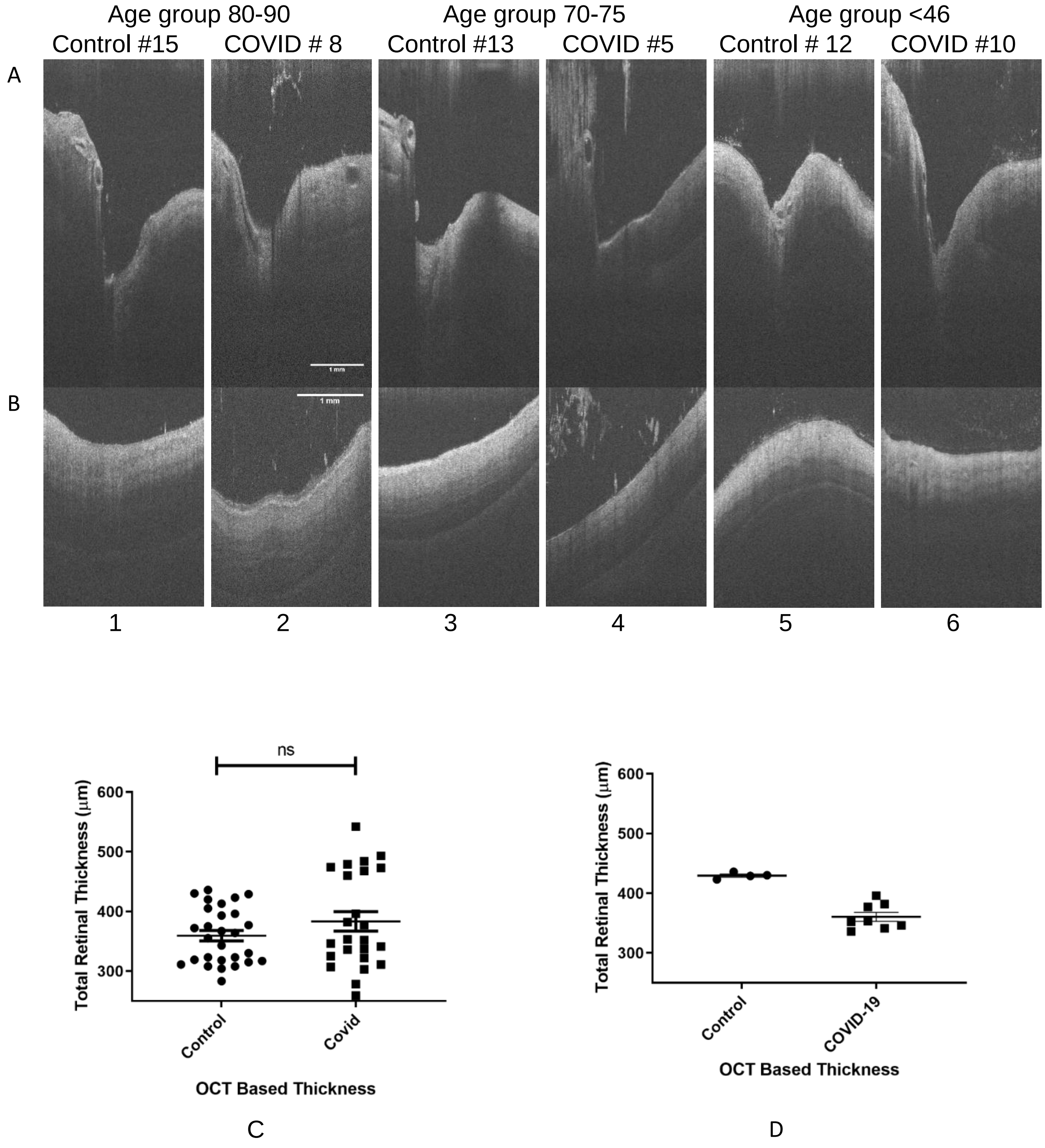
Age group 80-90
Age group 70-75
Age group <46
Control #15
COVID # 8
Control #13
COVID #5
Control # 12
COVID #10
A
B
1 mm
1
2
3
4
5
6
ns
Total Retinal Thickness (μm)
600
500
400
300
Control
Covid
OCT Based Thickness
C
Total Retinal Thickness (μm)
600
500
400
300
Control
COVID-19
OCT Based Thickness
D

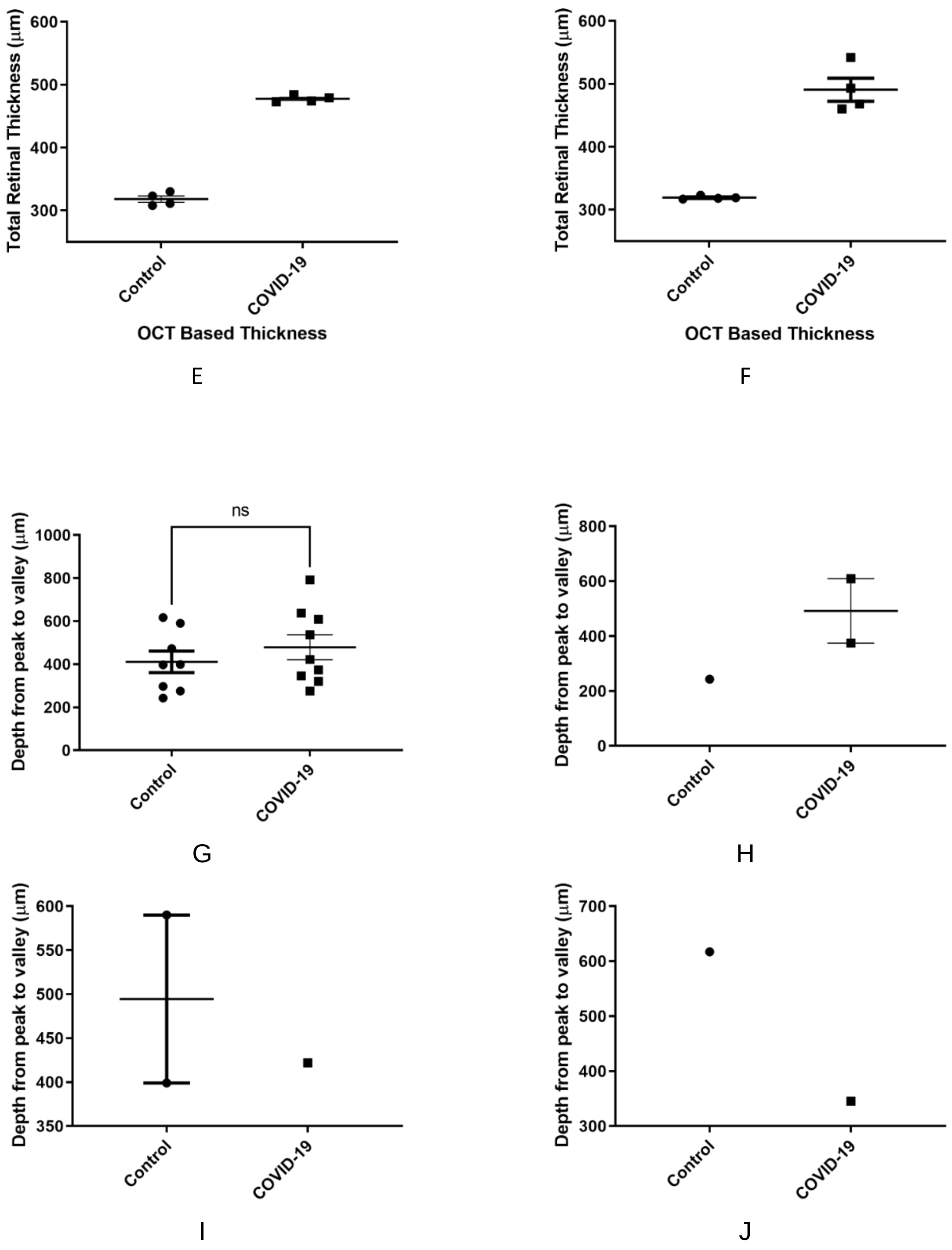


**Fig. S1:** Optical coherence tomography (OCT) cross sectional images of COVID-19 and age match control eyes, with a representative B-scan of (A) optic nerve head (ONH) and (B) central retina. The three age groups, 80-90, 70-75, and <46 were compared for retinal integrity and ONH cup size. On average, the ONH cup size was smaller in COVID-19 eyes in groups 80-90 and 70-75 when compared with control eyes. The retina layers integrity had no visible differences between the groups. Scale bar = 1 mm. Averaged total retinal thickness (RNFL to RPE) using OCT based retinal b-scans, with (C) All age groups, (D) age 80-90,

(E) age 70-75, and (F) age <46. The difference was not significant between the two groups in all age category, and statistical test was not performed on remaining groups because of less number of samples. The average total retinal thickness (measured at four location, each 200 µm apart, on the b-scan) was slightly increased in COVID-19 but was not statistically significant (i.e. $P<0.05$, in a two-tailed parametric t-test). Optic nerve head depth measurement (from peak to valley) using OCT images, with (G) All age groups (H) age group <46, (I) age group 70-75, and (J) age group 80-90. On average, the difference was not statically significant between the two groups in the all age group comparison (G).

**Figure S2**

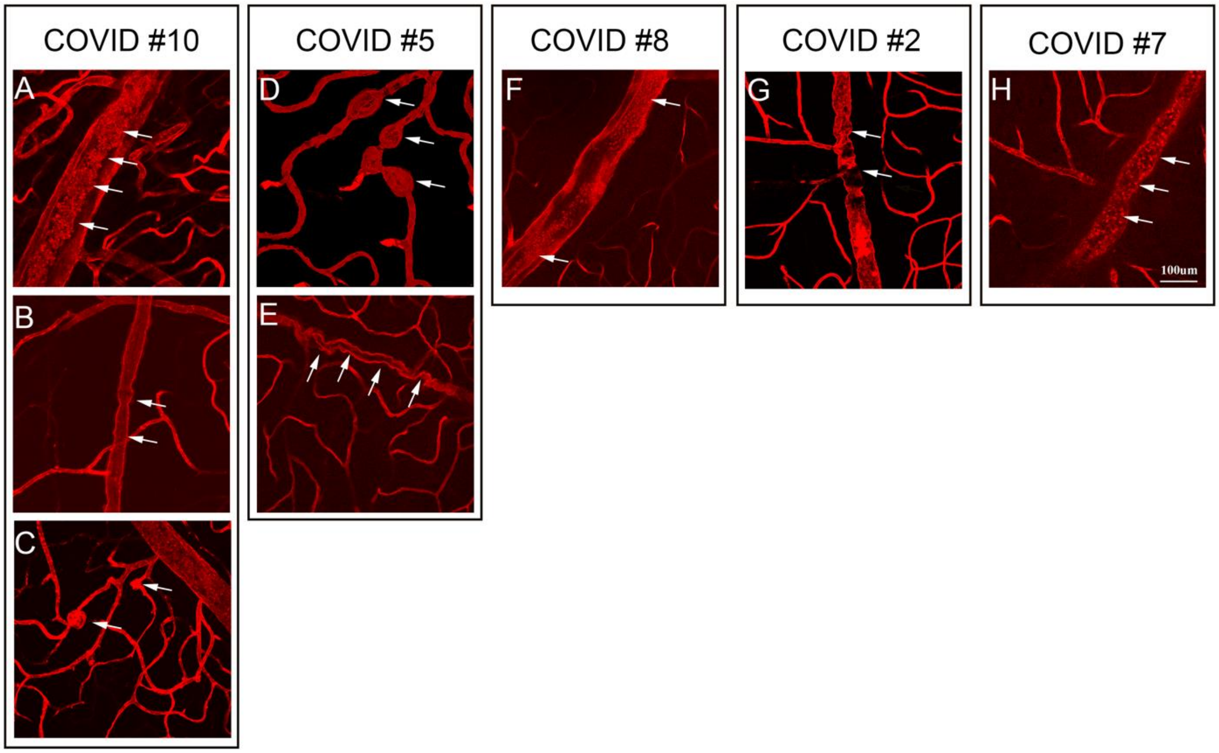


**Fig. S2:** Detection of other vascular abnormalities in COVID-19 patients after staining with lectin: (A, F, H) Representative images of the retinal vein occlusion, (B, E, G) vessels show marked tortuosity, (C, D) signifies vessel wall thickening which is indication of retinal arterial macroaneurysms in the COVID-19 patients. Scale bar:100 µm.

**Figure S3**

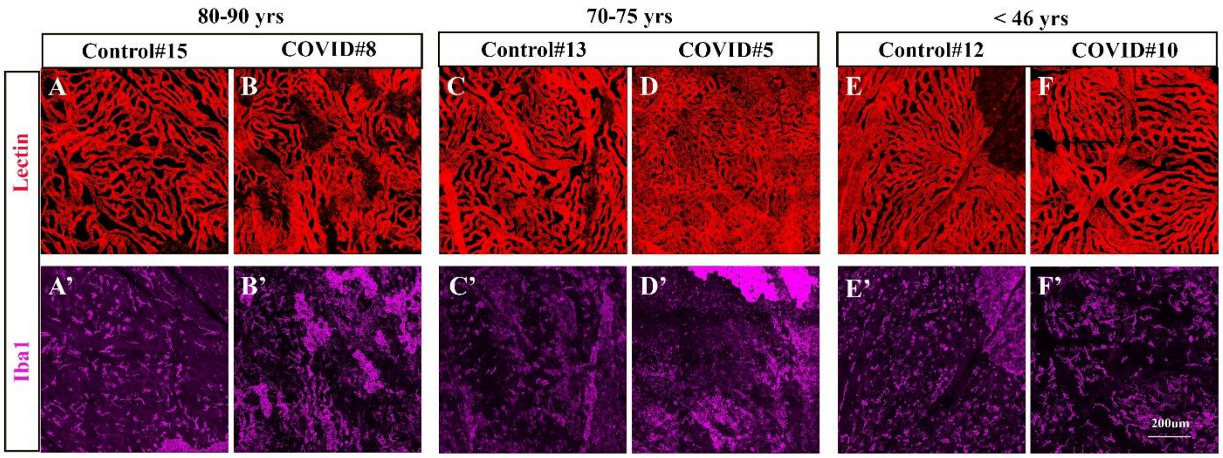


**Fig. S3:** Choroidal vasculature and microglial cells from COVID-19 and control cohorts (A-F'). Representative images (A-F) labeled with Rhodamine-UEA-Lectin (red) and (A'-F') Iba1 (Magenta). Choroidal vascular density appears to be similar in the COVID (B, D. F) and non-COVID individuals (A,C,E). Increase in Iba-1+ cells in the COVID-19 samples (B', D',F') was observed, although the increase is not consistent throughout the choroid. Scale bar = 200μm.
